# Enrichment of Rare Variants of Hemophagocytic Lymphohistiocytosis Genes in Systemic Juvenile Idiopathic Arthritis

**DOI:** 10.1101/2024.03.13.24304215

**Authors:** Mariana Correia Marques, Danielle Rubin, Emily Shuldiner, Mallika Datta, Elizabeth Schmitz, Gustavo Gutierrez Cruz, Andrew Patt, Elizabeth Bennett, Alexei Grom, Dirk Foell, Marco Gattorno, John Bohnsack, Rae S. M. Yeung, Sampath Prahalad, Elizabeth Mellins, Jordi Anton, Claudio Arnaldo Len, Sheila Oliveira, Patricia Woo, Seza Ozen, INCHARGE Consortium, Zuoming Deng, Michael J. Ombrello

## Abstract

**Objective:** To evaluate whether there is an enrichment of rare variants in familial hemophagocytic lymphohistiocytosis (HLH) genes and systemic juvenile idiopathic arthritis (sJIA) with or without macrophage activation syndrome (MAS).

**Methods:** Targeted sequencing of HLH genes (*LYST, PRF1, RAB27A, STX11, STXBP2, UNC13D*) was performed in sJIA subjects from an established cohort. Sequence data from control subjects were obtained *in silico* (dbGaP:phs000280.v8.p2). Rare variant association testing (RVT) was performed with sequence kernel association test (SKAT) package. Significance was defined as p<0.05 after 100,000 permutations.

**Results:** Sequencing data from 524 sJIA cases were jointly called and harmonized with exome-derived target data from 3000 controls. Quality control operations produced a set of 481 cases and 2924 ancestrally-matched control subjects. RVT of sJIA cases and controls revealed a significant association with rare protein-altering variants (minor allele frequency [MAF]<0.01) of *STXBP2* (p=0.020), and ultra-rare variants (MAF<0.001) of *STXBP2* (p=0.007) and *UNC13D* (p=0.045). A subanalysis of 32 cases with known MAS and 90 without revealed significant association of rare *UNC13D* variants (p=0.0047). Additionally, sJIA patients more often carried ≥2 HLH variants than did controls (p=0.007), driven largely by digenic combinations involving *LYST*.

**Conclusion:** We identified an enrichment of rare HLH variants in sJIA patients compared with healthy controls, driven by *STXBP2* and *UNC13D*. Biallelic variation in HLH genes was associated with sJIA, driven by *LYST*. Only *UNC13D* displayed enrichment in patients with MAS. This suggests that HLH variants may contribute to the pathophysiology of sJIA, even without MAS.

## Introduction

Systemic juvenile idiopathic arthritis (sJIA) is a severe inflammatory condition with onset in childhood. It is characterized by recurrent episodes of high-spiking fevers, a classic salmon-colored rash, arthritis, generalized lymphadenopathy and serositis.^1^ Up to a third of the cases are complicated by macrophage activation syndrome (MAS), a secondary form of hemophagocytic lymphohistiocytosis (HLH), with up to 40% of patients having subclinical forms of the disease.^2,3^ Previous small studies have identified an enrichment of variants in genes associated with familial forms of HLH in patients with sJIA and a history of MAS. However, whether these variants associate with sJIA in general has never been evaluated Therefore, we performed a large, targeted sequencing study to investigate the role of HLH variation in sJIA.

## Patients and Methods

### Samples and Sequencing

#### Study populations

We studied sJIA patients from the International Childhood Arthritis Genetics (INCHARGE) Consortium (Supplementary Table 1). The patients were diagnosed with sJIA by pediatric rheumatologists from 9 countries according to International League of Associations for Rheumatology (ILAR) criteria,^1^ as previously described.^4,5^ Referring rheumatologists confirmed if a subgroup of patients had ever had a history of macrophage activation syndrome by Ravelli criteria.^2^ Sequencing data from the Atherosclerosis Risk in Communities (ARIC) Cohort, obtained *in silico* from the database of Genotypes and Phenotypes (dbGaP; accession number phs000280.v8.p2), were utilized as population control data. This study was conducted as non-human subjects research, as determined by the Institutional Review Board of the National Institutes of Health.

**Table 1.** Combination of variants on sJIA patients with more than one rare qualifying HLH variant.

| **MAS** |  | **LYST** | **STX11** | **PRF1** | **RAB27A** | **UNC13D** | **STXBP2** |
| --- | --- | --- | --- | --- | --- | --- | --- |
| **Positive** | P1 | Q562H |  |  |  | V779M |  |
| **Positive** | P2 | N2971K |  |  | R184Q |  |  |
| **Negative** | P3 | Y875C | L9R |  |  |  |  |
| **Negative** | P4 | N2971K, Q562H |  |  |  |  |  |
| **Negative** | P5 | V3775M, Q562H |  |  |  |  |  |
| **Unknown** | P6 | Q562H |  |  |  | R1074W |  |
| **Unknown** | P7 |  |  | T496I |  |  | A211T |
| **Unknown** | P8 | R3509Q, N2971K |  |  |  |  |  |
| **Unknown** | P9 | N3158S |  |  |  | H224Y |  |
| **Unknown** | P10 | R1718Q |  |  |  |  | P573L |
| **Unknown** | P11 |  | V197M | P501S |  |  |  |
| **Unknown** | P12 | R2624W, K1899N |  |  |  |  |  |
| **Unknown** | P13 | N2971K |  |  |  | R1074W |  |

Rare qualifying variants defined as minor allele frequency < 0.01, exonic, non-synonymous, passing quality filters, present in the canonical transcript.

#### Sequencing

The sJIA patients underwent targeted sequencing of genes associated with familial forms of HLH (*LYST, PRF1, RAB27A, STX11, STXBP2, UNC13D*) using custom Illumina Nextera Capture Assays. Libraries were sequenced via Illumina HiSeq or MiSeq sequencers. Population controls had undergone whole exome sequencing at the Human Genome Sequencing Center at the Baylor College of Medicine using Illumina HiSeq 2000 platform.^6^ Data from the target region were extracted from the whole exome sequencing (Supplementary Data 1).

#### Variant Calling

All cases and controls were jointly called using a variant discovery pipeline modelled after the Genome Analysis Toolkit (GATK 4.0.12.0) best practices (Broad Institute), and variants were filtered to only include high-quality variants that were well covered on both cases and controls. Details of variant-level quality control are described in the supplementary methods.

#### Exclusion of genetic outliers and related individuals

Related individuals were identified and removed with PLINK (v1.9) and KING (v2.2). We integrated data from HapMap3 reference populations and principal components analysis (PCA) was performed using SNP & Variation Suite 8.3.0 (Golden Helix, Bozeman, MT) to identify and remove ancestral outliers. See supplementary methods for full details.

### Rare Variant Association Testing

#### Selection of qualifying variants

Minor allele frequency (MAF) and variant function filters were applied to include rare (MAF <0.01), exonic, protein altering variants that mapped to the canonical Ensembl GRCh37.p13 transcript (Supplementary Table 2). As a sensitivity analysis, low-frequency (MAF < 0.05) or ultra-rare (MAF < 0.001) variant sets were also examined.

#### Statistical tests

Rare variant association testing (RVT) was performed in R (version 4.3.2) using the Sequence kernel association test package (SKAT, version 2.2.5). The primary RVT was performed at the gene and group level using the unweighted SKAT with 100,000 permutations.^7^ SKAT is a variance-component test, which enables comparison of variant distributions between 2 groups with the least assumptions. As a sensitivity analysis, we used the SKAT package to perform burden testing. Significance was evaluated at a threshold of P_permutation_ ≤ 0.05. Frequency of individuals with 2 or more HLH variants was analyzed using two-tailed Fisher’s exact test in R.

## Results

Targeted sequencing was performed in 525 sJIA patients and sequencing data from the target region were extracted from 3000 population control subjects. Joint variant calling of the 3525 samples revealed data corruption in 1 sJIA case, which was excluded. Subjects with dissimilar genetic ancestry were excluded (39 cases, 22 controls), as were those with cryptic relatedness (48 controls) or data missingness over 10% (6 controls). Additionally, 4 sJIA cases were excluded after being found to have monogenic autoinflammatory disease. This produced a final study population of 481 sJIA patients and 2924 control subjects (Supplementary Figure 1). After joint calling, the dataset included 622 variants in 6 HLH genes. Coverage harmonization revealed 115 variants had unbalanced coverage between cases and controls, which were excluded (Supplementary Data 2). Eight additional variants did not meet the hard variant quality control thresholds and were excluded. The final dataset included 499 variants (Supplementary Figure 2, Supplementary Data 3).

Among these 499 variants, 367 were observed in control subjects and 161 were observed in sJIA cases. The sJIA variants included 64 synonymous variants, 5 variants of the 5’ or 3’ untranslated region and 92 protein-altering variants (91 nonsynonymous variants and 1 non-frameshift deletion). None of the HLH variants observed in cases were classified as pathogenic by ClinVar, whereas 11 variants in the control population were pathogenic, 2 of which were excluded for unbalanced coverage (Supplementary Data 2 and 3). Three of the variants observed in sJIA cases were not present in the gnomAD database. The first (*UNC13D* E1066D) was observed in a patient with known MAS, while the other 2 (*LYST* Q2628R and *LYST* K1729M) were observed in patients with unknown MAS status. There were no rare variant homozygotes among the sJIA patients, however three sJIA patients did carry homozygous, low-frequency variants.

These included *UNC13D* R928C, which is listed as benign/likely benign on ClinVar; *PRF1* A91V, which has been shown to cause partial cytolytic dysfunction but is also often observed in healthy individuals,^3^ and *UNC13D* A59T, which has been observed in homozygosity in a healthy individual without HLH.^8^ Finally, another sJIA case who was excluded for dissimilar ancestry was homozygous for *UNC13D* A59T, while also carrying two *LYST* variants (D2228H and N1228S).

### Rare/ultra-rare coding variants of *STXBP2* and *UNC13D* are associated with sJIA

In the primary subgroup of variants (MAF < 0.01), 481 sJIA cases carried 110 alternate alleles of 71 unique HLH gene variants, and 2924 control subjects carried 517 alleles of 198 variants (Supplementary Table 3). Visual comparison of the aggregated alternate allele frequencies of HLH genes revealed a higher frequency of rare, protein-altering variants in sJIA cases than in controls in each comparison (Figure 1). To evaluate whether these differences reflected a significant difference in the distribution of variants, we performed RVT of rare variants of HLH genes by SKAT (Supplementary Table 4).

**Figure 1.**
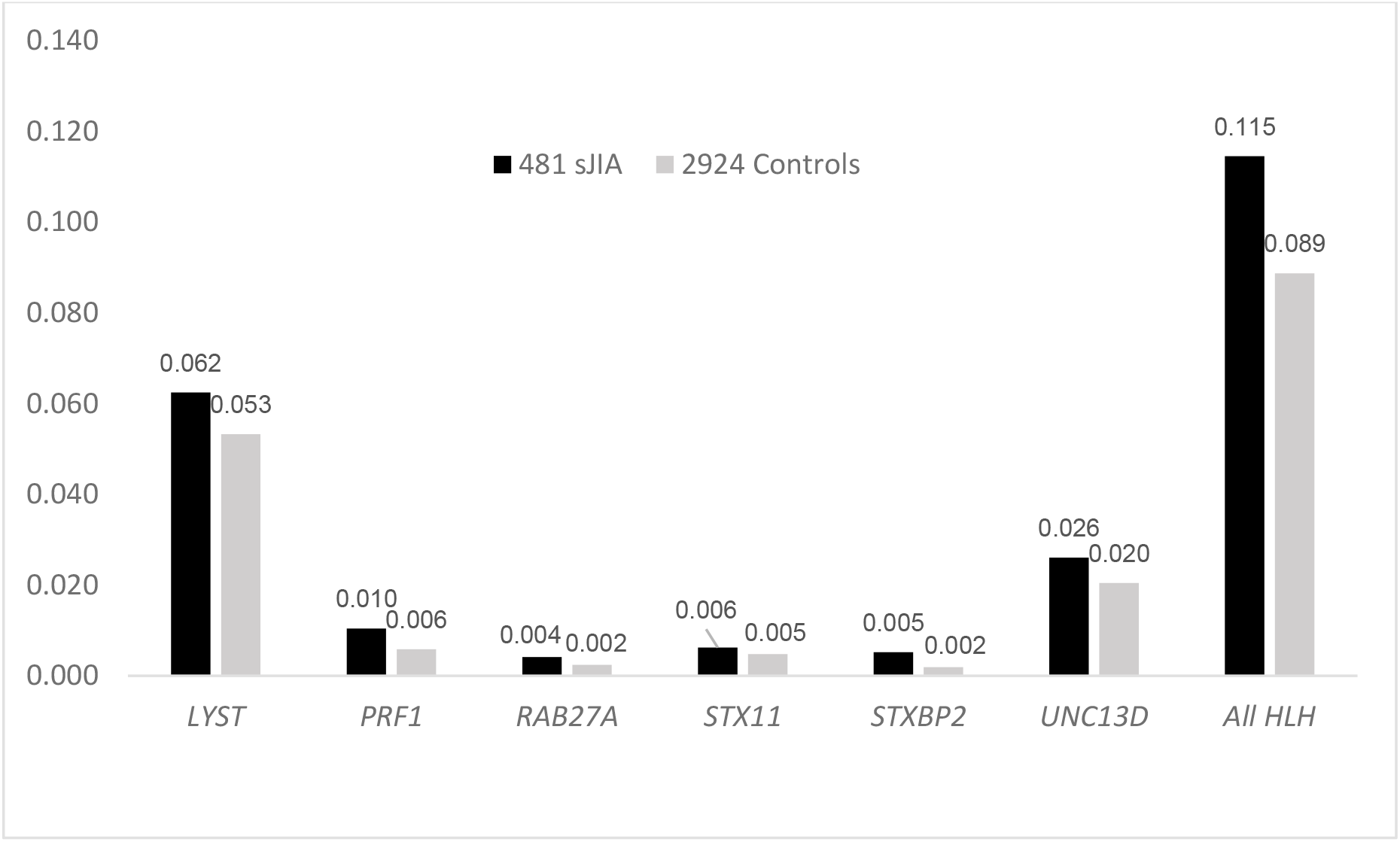
Aggregated allele frequencies from crude counts of rare qualifying variants in the sJIA cohort compared to controls. Aggregated alternate allele frequencies were calculated as the total count of alternate alleles in all the variants in the gene, divided by the mean number of alleles identified in all the variants in the gene (to account for missing genotypes). Rare qualifying variants defined as minor allele frequency <0.01, exonic, non-synonymous, passing quality filters, present in the canonical transcript. Burden test showed significant difference in all HLH genes as a panel (p = 0.014) and SKAT showed significant association of *STXBP2* (p= 0.020).

This revealed a significant association between sJIA and *STXBP2* (p = 0.020). When SKAT was repeated on the set of ultra-rare variants (MAF < 0.001), the association of *STXBP2* was even more significant (p = 6.7E-3), an association with *UNC13D* variation was also identified (p = 0.045), and the HLH genes as a panel became significant (p = 0.011). We observed no significant association of sJIA with low frequency (MAF < 0.05) variants of HLH genes.

### Development of MAS in JIA is associated with rare, protein-altering variants of *UNC13D*

To evaluate the relationship between rare HLH gene variants and MAS, we examined 122 sJIA cases in our cohort who were known to have either had MAS (MAS+; n = 32) or to have never had MAS (MAS-; n = 90) by Ravelli criteria.^2^ (Supplementary Tables 5, 6, 7 and 8). Association testing with SKAT identified associations between MAS+ and *UNC13D* variation in comparison with either MAS- or healthy controls (p = 0.0047 and p = 0.038, respectively).

### Full sJIA cohort and MAS+ cases have enrichment of rare, protein-altering variants of HLH genes

In situations where rare variants are expected to have the same directional effect on disease risk, burden testing provides greater statistical power to detect rare variant associations than SKAT. Given that HLH-causing variation collectively produces HLH through a reduction of cytolytic capacity,^3^ we employed burden testing as a sensitivity analysis (Supplementary Tables 4 and 6). This revealed that rare, protein-altering variants of HLH genes as a panel were significantly enriched in children with sJIA, relative to control subjects (p = 0.014), and despite the smaller sample size, the same association was seen in MAS+ compared to controls (p = 0.030), but not on MAS- (p = 0.151). Rare variants of *PRF1* were significantly enriched in MAS-relative to controls by the burden test (p = 0.029), but not in the primary SKAT analysis.

Additionally, sJIA cases more often carried 2 or more rare, protein-altering HLH variants (13/481, 2.7%) than did control subjects (31/2924, 1.1%; p = 0.007, OR 2.6 [1.2, 5.1]; Figure 2). Although the MAS+ and MAS-patients carried 2 or more HLH variants at rates higher than the sJIA or control cohorts (2/32, 6.3% and 3/90, 3.3%, respectively), the differences were not statistically significant.

**Figure 2.**
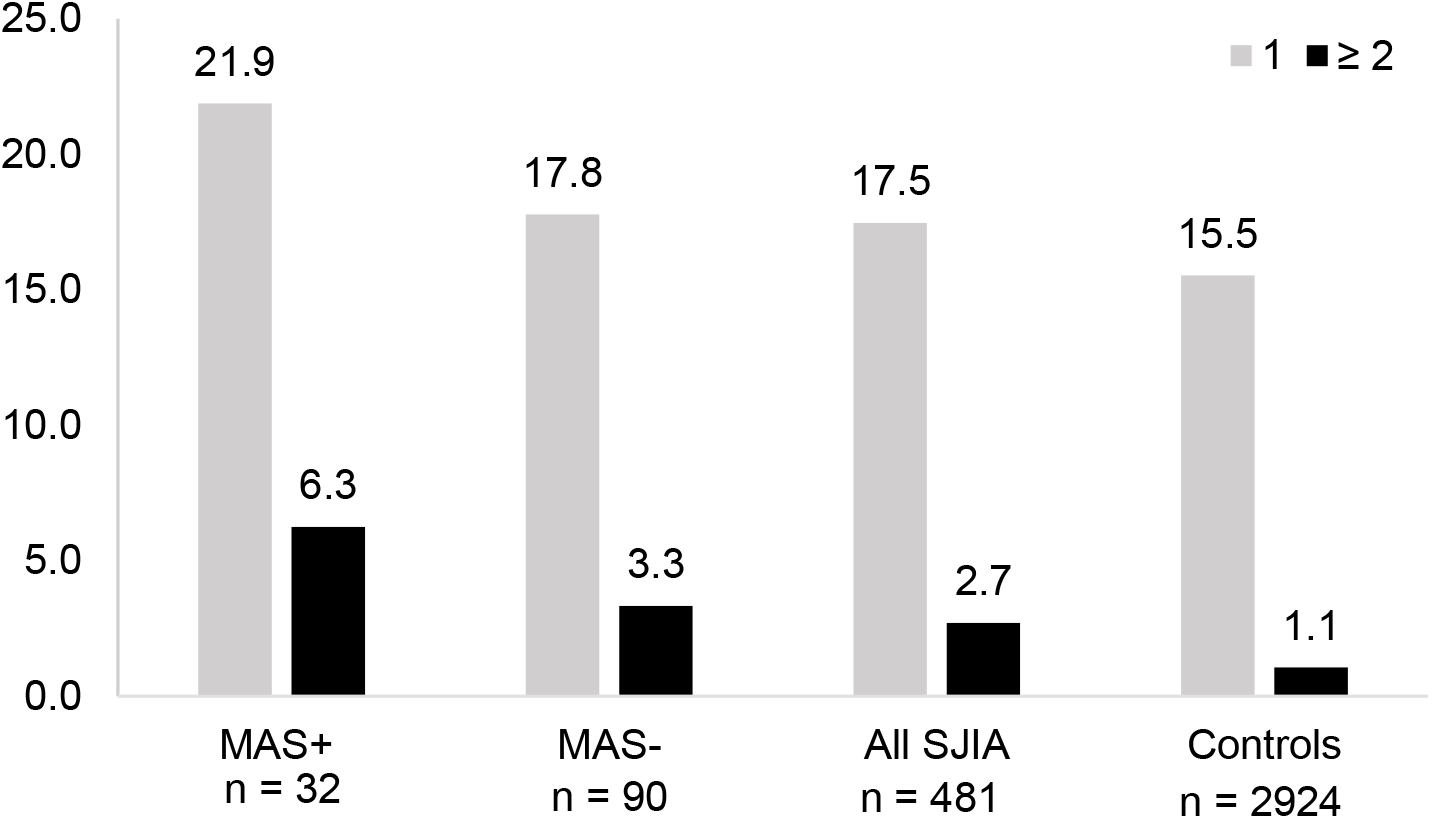
Distribution of individuals with one or more rare, protein-altering HLH variants. A bar plot displays the proportion of patients with either 1 or ≥2 rare (MAF < 0.01), protein-altering variants on canonical HLH gene transcripts among sJIA patients with a history of MAS (MAS+), sJIA patients with no history of MAS (MAS-), the full sJIA cohort and the control collection. MAS, macrophage activation syndrome. MAF, minor allele frequency. HLH, hemophagocytic lymphohistiocytosis.

Variation in HLH genes causes congenital HLH in a recessive fashion, but it has also been suggested variants in different HLH genes could synergize to cause pathology in a digenic or oligogenic fashion.^9^ We examined the set of variants that were observed in these combinations (Table 1, Supplementary Data 4) and observed that most combinations involved *LYST* in both cases and controls. However combinations involving *LYST* occurred significantly more commonly in sJIA (11/481, 2.2%) than in controls (24/2924, 0.8%; p = 0.007; OR 2.8 [1.2,6.0]). While sJIA patients also more often carried biallelic *LYST* variation than did controls (0.8% vs. 0.2%), the difference was not statistically significant.

## Discussion

This study identified an association of rare/ultra-rare variants of *STXBP2* and *UNC13D* in sJIA patients compared to controls. The association with *UNC13D* seems to have been indeed driven by the patients with MAS in the cohort, since the association was apparent in the subset of patients with a known history of MAS, despite the smaller size of this cohort. This finding aligns with previous studies that had identified an increased burden of HLH variants as a whole and an enrichment of *UNC13D* variants in sJIA patients with MAS.^3^ In contrast, the *STXBP2* association was only found in the full sJIA cohort, and it was not identified in the MAS positive patients. This could have been due to a lack of power, given that the cohort of MAS patients was much smaller than the full sJIA cohort. However, the smaller size did not limit the identification of the *UNC13D* association, so it is possible that the association of *STXBP2* is truly being driven by the patients without MAS in the cohort, and it is playing a role in the pathophysiology of sJIA itself. *STXBP2* variants have been previously described in sJIA patients with MAS,^3^ but the smaller sample size of the study did not allow for evaluation of associations of individual genes.

This study identified that sJIA patients in general and those with MAS have an increased burden of rare HLH variants as a panel compared to controls, which was even more pronounced in the ultra-rare MAF cutoffs. Furthermore, sJIA patients more often carried 2 or more HLH variants than controls, which was driven by *LYST* variants in combination with variants in other genes (or in *LYST*). It is notable that although *LYST* was not associated with sJIA by itself, its presence in combination with another HLH variant was significantly associated. This is consistent with the hypothesis that variants in *LYST*, which is involved in granule biosynthesis and generally leads to milder cytolytic defects, may interact with heterozygous variants in terminal granule processing genes and produce pathology.^9^

Similar to a previous study of MAS in lupus^10^ and unlike a study of secondary HLH in adults,^11^ we did not identify significant associations of low-frequency HLH variants in sJIA or MAS. The association of rare variants as a panel was only identified by burden tests, which could be attributable to its higher power to identify associations when all variants have the same directionality of effect. In this way, it would also be more sensitive to identify synergistic effects of combined hypomorphic variants. We did not identify a significant association of HLH variants in the MAS-group compared to controls in the primary analysis. However, we did identify an enrichment of rare *PRF1* variants in this group by burden test. Therefore, it is possible that in larger studies of sJIA patients without MAS, an association with *PRF1* could rise to significance.

This study did not recapitulate previous findings of an association of *PRF1* variants in patients with MAS.^12^ Specifically *PRF1* A91V, which had been previously associated with MAS in sJIA,^12^ and with severity of secondary HLH,^11^ was more common in sJIA patients without MAS than in patients with MAS in our cohort and had similar incidence in sJIA patients and controls. Functional studies of this variant have shown it to cause a partial loss of protein function and stability, however, it has been vastly reported in unaffected individuals as well.^3^ We did identify one patient of unknown MAS status who was homozygous for *PRF1* A91V, which has been shown to cause transient cytolytic defects in the setting of HLH.^13^ Our study helps to increase the understanding that even though *PRF1* A91V variants might predispose to HLH in the context of infections^13,14^ or immune reconstitution syndrome,^15^ this variant does not seem to confer an increased risk of development of MAS in the inflammatory background of sJIA. This could suggest that the increased susceptibility to HLH in the context of infection could be related to a component of immunodeficiency.

A major strength of this study is its power. As the largest examination of HLH gene variation in sJIA to date, it provides the best estimate of the burden of HLH variation among patients with sJIA and the first population-level assessment of these genes on an individual basis. Another advantage of this study design was its use of a large population control dataset. This enabled the study to move beyond investigations of sJIA/MAS and discover new relationships between HLH gene variants and sJIA, itself. Furthermore, the analysis of individual sequencing data allowed to compare the frequency of digenic HLH variants in cases and controls, as well as the combination of variants.

This study also had some limitations. We did not have detailed clinical data or functional studies from the patients or controls included in this study, so it was not possible to determine if their variants had functional effects. Furthermore, due to lack of parental data, we could not determine if multiple variants were inherited in *cis* or *trans*. The INCHARGE cohort is largely composed of individuals of European descent, and it was necessary to create an ancestrally focused study population to minimize the potential effects of population stratification. As a result, it will be important to perform similar studies in patients from other ancestries. Although our variants were not Sanger validated, we implemented strict quality cutoffs for variant selection to avoid the inclusion of false positive calls.

Future studies of large cohorts of sJIA patients from diverse backgrounds will allow for the verification of these findings in patients with different ancestries. Ongoing efforts to assemble large cohorts with detailed clinical datasets that can be linked to sequencing data and biospecimens will help clarify further the associations of HLH variants in sJIA patients without MAS.

## Data Availability

"All data produced in the present study are available upon reasonable request to the authors."

## Acknowledgements

This study was funded by the Intramural Research Program of the National Institute of Arthritis and Musculoskeletal and Skin Diseases (Z01-AR041198). The work utilized the computational resources of the NIH high-performance computing cluster Biowulf (http://hpc.nih.gov)

Dr. Prahalad is supported in part by The Marcus Foundation Inc., Atlanta.

“The Atherosclerosis Risk in Communities study has been funded in whole or in part with Federal funds from the National Heart, Lung, and Blood Institute, National Institute of Health, Department of Health and Human Services, under contract numbers (HHSN268201700001I, HHSN268201700002I, HHSN268201700003I, HHSN268201700004I, and HHSN268201700005I). The authors thank the staff and participants of the ARIC study for their important contributions.”

